# Protocol for the REVELIO test-track pilot study: a randomised, controlled, single-centre trial in healthy recreational cannabis users investigating real-time in-vehicle detection of cannabis-impaired driving

**DOI:** 10.64898/2026.04.29.26352110

**Authors:** Robin Deuber, Michal Bechny, Christoph Heck, Joel Brügger, Matthias Pfäffli, Mia Jovanova, Elgar Fleisch, Felix Wortmann, Wolfgang Weinmann

## Abstract

**Background:** Driving under the influence of cannabis is associated with impaired cognitive and psychomotor performance and an increased risk of traffic accidents. Reliable real-time in-vehicle systems for detecting cannabis-related driving impairment are currently lacking; but hold great potential for improving road safety.

**Methods:** This protocol describes the REVELIO test-track pilot study: a randomised, controlled, open-label, interventional, single-centre trial. The study assesses the feasibility and methodological requirements for developing and evaluating a multimodal in-vehicle detection approach using vehicle and driver state data. A total of 45 healthy recreational cannabis users will be enrolled and randomly allocated to an intervention or a reference control group. During the main study day, all participants will undergo biological sampling for tetrahydrocannabinol (THC) and related metabolites, as well as pre-driving assessments, followed by a sober baseline driving session on a closed test track using a dual-pedal vehicle with a certified driving instructor onboard. Participants in the intervention group will then receive a single controlled inhalative cannabis dose (target 0.67 mg THC per kg body weight), while the reference group will receive no cannabis. All participants will subsequently complete three additional standardized 50-minute driving sessions at predefined time points up to approximately six hours after administration, following identical schedules to enable within- and between-group comparisons. Between driving sessions, structured breaks will include recovery periods, repeated biological sampling, and traffic-medical, traffic-psychological, and pre-driving performance assessments, to characterise the temporal dynamics of cannabis-related impairment.

**Discussion:** Multimodal data will be collected, including vehicle controller area network (CAN) data, driver monitoring camera (DMC) data, physiological signals using wearables, and biological samples (capillary blood, breath, oral fluid.

Machine-learning–based models will be developed and evaluated to distinguish sober from cannabis-influenced driving states under controlled conditions. Secondary analyses will examine changes in driving performance over time and associations between functional measures and biological THC concentrations. As an exploratory pilot study conducted on a secured test track, the protocol aims to generate standardized reference data and quantitative performance metrics to inform both feasibility and system design considerations.

**Ethics and trial registration:** The study was approved by the Cantonal Ethics Committee Bern, Switzerland (BASEC ID: 2025-01590) and is registered at ClinicalTrials.gov (NCT07401628).

## Introduction

Drug-impaired driving represents a significant and growing global road safety challenge. In the United States, 56% of seriously or fatally injured road users tested positive for at least one drug, including alcohol. Cannabinoids, specifically tetrahydrocannabinol (THC), was the most frequently detected illicit substance (25.1% of tested cases) [1].

European data show a comparable pattern, identifying THC as the most prevalent illicit drug among drivers, with a weighted average prevalence of 1.3% across 13 countries and country-specific rates of up to 6% [2].

Evidence from epidemiological studies consistently links acute cannabis consumption to an increased risk of motor vehicle collisions and measurable impairments in driving performance [3–5]. Population-level analyses further suggest that legalization and commercialization of recreational cannabis are associated with increases in traffic injury and fatality rates [6–8], and continued expansion of legal cannabis markets is expected to further increase the prevalence of cannabis use among drivers [9].

Cannabis consumption impairs a broad range of cognitive, psychomotor, and perceptual functions that are essential for safe driving [9, 10]. The most consistently affected cognitive domains include visual and divided attention, reaction time, motor tracking, executive control, and time perception [11–15]. At the driving-behavioral level, studies report impaired vehicle control, reflected by increased lateral position variability and lane deviation, reduced cornering stability, increased braking distance, and delayed responses to traffic signals [14, 16–21]. In contrast to alcohol, cannabis-related functional driving impairment appears more heterogeneous across individuals.

Consistent with this variability, several studies report no systematic changes in speed- or steering-related metrics under cannabis influence [14, 15, 19, 22, 23].

This variability is influenced by inter-individual differences and may be partly attributable to dose, cannabis composition (e.g., THC content and formulation), route of administration, prior exposure, tolerance, and complex pharmacokinetics, including variability in absorption, distribution, and elimination [24–26].

Current approaches for detecting driving under the influence rely primarily on roadside testing or laboratory analysis of biological samples and therefore assess substance presence rather than functional driving performance at the time of vehicle operation [1, 2, 27]. These approaches are typically applied retrospectively, for example during police roadside checks or following traffic accidents, and hence do not enable continuous or real-time assessment of driving capability. In contrast, existing in-vehicle systems for monitoring driver states such as drowsiness and distraction typically rely on observable behavioral indicators (e.g., eye and head movements) and driving patterns extracted from vehicle sensor data [28–31].

Research on alcohol-impaired driving detection has shown that impairment can be reliably inferred from driving patterns and driver state indicators [32–38]. In controlled and experimental settings, machine learning approaches have demonstrated robust discriminative performance, frequently achieving area under the receiver operating characteristic curve (AUROC) values *≥*0.80 and accuracies exceeding 80-90%, depending on study design and data modality. Crucially, these approaches are anchored in legally defined blood- and breath-alcohol concentration thresholds (e.g., 0.05%) that exhibit a comparatively stable and dose-dependent relationship with functional driving impairment [39, 40]. At the same time, this body of work has underscored methodological challenges related to inter-individual variability, generalisation to unseen drivers, and false alarm control—issues that are directly relevant to the development of cannabis-impaired driving detection systems.

For cannabis, however, concentration-based detection approaches, including zero-tolerance and threshold-based (*per se*) policies relying on biological sampling (c.f. [41–44]), are challenged by complex pharmacokinetics, substantial inter-individual variability and an inconsistent relationship between biological THC concentrations and functional driving ability [24, 25]. To date, despite extensive evidence of cannabis-related impairment of driving-relevant skills [9–21], *no scalable, real-time, in-vehicle detection system for cannabis-impaired driving has yet been developed or systematically evaluated*. Existing research has primarily focused on characterising impairment effects retrospectively rather than on assessing whether driving-related data can support real-time detection during vehicle operation.

Motivated by this gap, the REVELIO test-track pilot study was designed to generate standardized, multimodal reference data for in-vehicle cannabis-related impairment detection under controlled and safety-oriented conditions. Using a randomised, controlled design, healthy recreational cannabis users will be assigned to an intervention or reference control group, enabling within-subject comparisons between cannabis-influenced and sober driving as well as between-group comparisons under identical driving conditions. The primary aim is to assess to what extent in-vehicle sensor data, including CAN signals, DMC data, and physiological signals, can be used to detect cannabis-impaired driving using machine-learning–based models. Secondary aims include comparing driving performance across different states of cannabis impairment over time, characterising changes in driving behaviour, driver state, and biological measures, and evaluating the relationship between functional driving performance and THC concentrations measured in capillary blood, breath, and oral fluid. Beyond cannabis-specific detection, the REVELIO dataset is intended to support subsequent methodological work on a cause-independent “fit-to-drive” model—that is, a model designed to detect functional driving impairment irrespective of its underlying cause (e.g., cannabis, alcohol, or metabolic disturbances). This approach will be informed by pooled analyses of datasets from prior experimental driving studies conducted by the same research group, including alcohol- and hypoglycaemia-related impairment, all of which followed closely aligned study protocols and comparable data acquisition procedures (ClinicalTrials.gov IDs: NCT04035993, NCT04980846, NCT05796609).

## Materials and methods

This study will be conducted in accordance with ethics study plan, version 5.1, approved by the responsible ethics committee, and available as Supporting File S2.

### Study design and setting

The REVELIO test-track pilot study is a randomised, controlled, open-label, interventional, single-centre trial using a parallel-group design with an allocation ratio of approximately 3:1, anticipating enrolment of 33 participants in the intervention group and 12 in the reference group. The trial follows an exploratory pilot framework designed to assess feasibility and diagnostic performance of machine-learning–based real-time detection of cannabis-impaired driving using multimodal in-vehicle sensor data under controlled and safety-oriented conditions. The primary aim is to evaluate to what extent driving-related sensor data, collected in real vehicles, can distinguish sober driving from time-resolved levels of cannabis influence corresponding to early, intermediate, and late post-consumption phases. As an exploratory pilot study in the absence of an established reference system, the trial is designed to assess feasibility and the diagnostic (classification) performance of distinguishing sober driving from defined post-consumption phases.

The study is conducted as a single-centre investigation coordinated by the Institute for Forensic Medicine (Institute für Rechtsmedizin, IRM), University of Bern, Switzerland, in collaboration with the ETH Zürich, Switzerland and the University of St. Gallen, Switzerland. Detailed overview of the schedule of study enrolment, interventions, and assessments is depicted in Fig. 1, adhering to updated SPIRIT (Standard Protocol Items: Recommendations for Interventional Trials) guidelines [45]. The Supporting File S1 additionally provides detailed SPIRIT 2025 checklist. The study comprises telephone screening and two on-site visits: a *presence screening* (Visit 1) conducted at the IRM, and a full-day *driving assessment* (main study day; Visit 2) performed on a secured, closed test track in Thun, Switzerland. The test track is not part of the public road network and is exclusively accessible to the study team during study days, ensuring a standardized and risk-minimising driving environment. All driving sessions are carried out in a dual-pedal study vehicle (2020 Volkswagen Touran 1.5 TSI, 110 kW, automatic transmission, 7-seat configuration) with a certified driving instructor seated in the front passenger seat, allowing immediate intervention if required.

**Fig 1.**
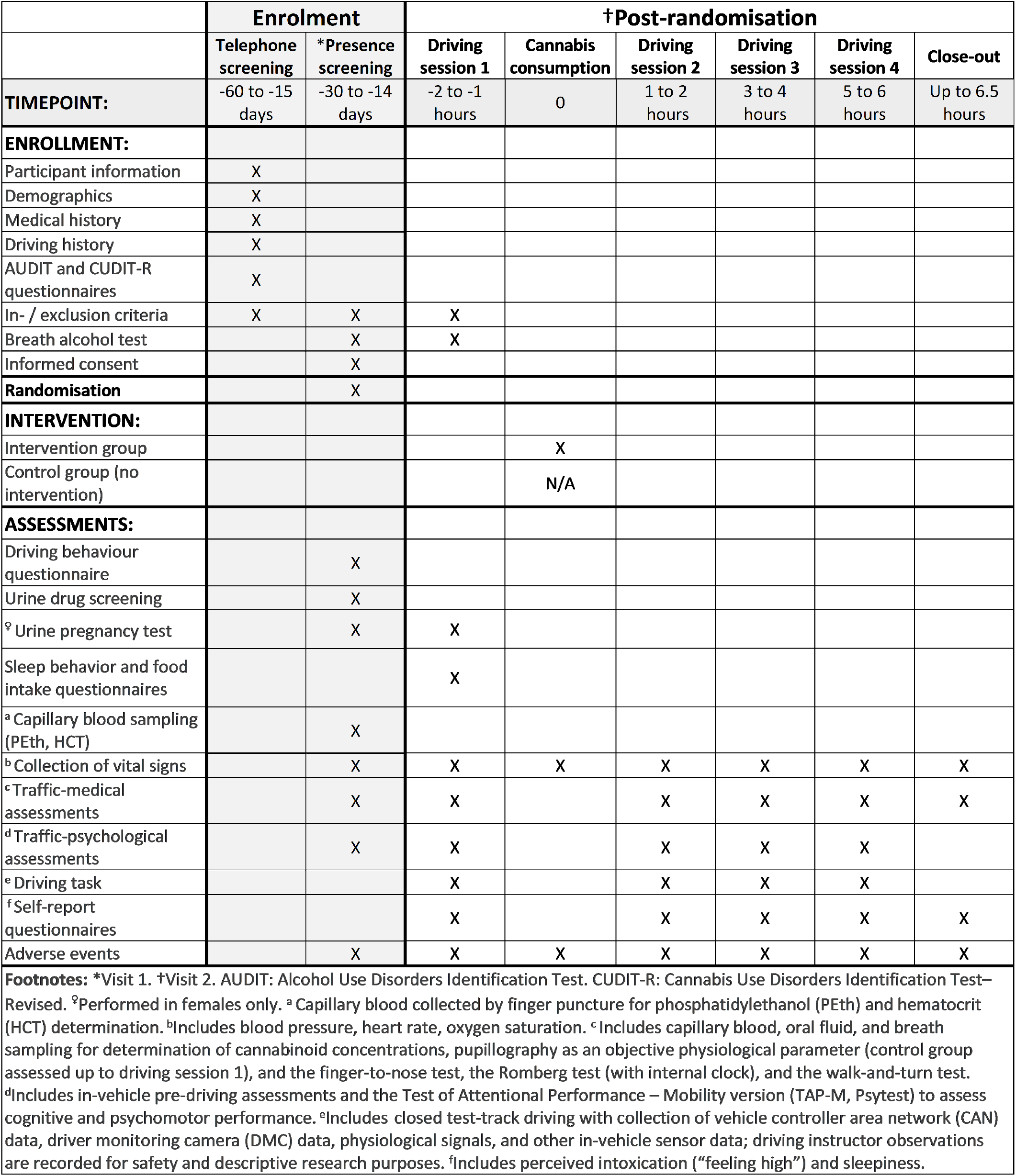
SPIRIT 2025 diagram of the schedule of enrolment, interventions, and assessments within REVELIO study.

Using a randomised, controlled, parallel design, healthy recreational cannabis users (see eligibility criteria) are allocated to either an intervention group or a reference control group. All participants first complete a sober baseline driving session.

Subsequently, participants in the intervention group receive a single, controlled inhalative cannabis administration (see intervention description), whereas participants in the reference group undergo identical study procedures without cannabis exposure.

After cannabis administration in the intervention group, all participants complete three additional standardized driving sessions conducted according to an identical schedule in both groups. Intervals between driving sessions are used for structured recovery periods, repeated biological sampling, and traffic-medical, traffic-psychological, and pre-driving performance assessments to capture temporal changes in driving-related function and biomarker dynamics.

The non-exposed reference group provides essential negative-labeled control data for the development and evaluation of machine-learning–based detection models, including estimation of false positive rates and assessment of generalisability. Beyond within-subject comparisons to each participant’s sober baseline, the reference group enables between-group analyses that help disentangle cannabis-associated effects from time-dependent influences, such as learning or fatigue, that may occur across repeated driving sessions even in the absence of cannabis exposure.

The study is designed as a pilot investigation to generate standardized multimodal data and to examine methodological, technical, and analytical aspects relevant to in-vehicle detection of cannabis-impaired driving. The focus is on feasibility, data quality, and performance characteristics, providing a structured basis for subsequent development and validation studies.

### Participants

A total of 45 healthy adult recreational cannabis users with regular driving experience will be enrolled. Participants will be randomly allocated to either a cannabis intervention group (n = 33) or a non-exposed reference group (n = 12).

#### Eligibility criteria

Eligibility criteria are predefined in the ethics-approved protocol and are assessed during telephone screening and both on-site visits (Visit 1 and Visit 2), as applicable. No vulnerable populations are included and all participants are healthy volunteers.

Inclusion criteria require that participants:

- Provide written informed consent prior to any study-specific procedures;
- Are recreational cannabis consumers (use more than once per month);
- Hold a valid Swiss or EU driving license, with a probationary period elapsed;
- Are 21 years of age or older;
- Have actively and regularly driven a car within the last 6 months;
- Are in good general health;
- Require no special equipment for driving (e.g. adaptive seats or controls);
- Are fluent in (Swiss) German and have no speech impairment. Exclusion criteria include:
- Medical conditions for which cannabis consumption is contraindicated, including cardiovascular disease, psychiatric or neurodevelopmental conditions (e.g., psychosis, depressive disorders, or attention-deficit/hyperactivity disorder), neurological or neuro-degenerative diseases, or other clinically relevant conditions;
- Cannabis abstinence;
- Excessive cannabis use, assessed using the Cannabis Use Disorders Identification Test–Revised (CUDIT-R) questionnaire [46] (question 1: 0 points, or sum *>*7 points);
- Pregnancy, breastfeeding, or intention to become pregnant during the study period;
- Alcohol misuse or risky drinking behaviour, defined by an Alcohol Use Disorders Identification Test (AUDIT) [47] score *≥*8 or phosphatidylethanol (PEth) concentration in capillary blood *>* 200 ng/mL at Visit 1;
- Positive breath alcohol test at Visit 1 or Visit 2;
- Use of drugs of abuse other than cannabis within 4 weeks prior to the study;
- Use of medications that may interfere driving ability;
- Inability to comply with study procedures due to language barriers, cognitive impairment, or psychological disorders;
- Body mass index *<* 18.5 kg*/*m^2^ or *>* 30 kg*/*m^2^;
- Participation in another clinical trial that may interfere with the present study according to the responsible investigator(s);
- Any relationship of dependency (hierarchical or social) with study personnel.

#### Sample size

The primary objective of this study is to assess the feasibility of machine-learning–based detection of cannabis-impaired driving using in-vehicle sensor data. Conventional power-based sample size calculations are not directly applicable to such detection tasks. Sample size planning was therefore informed by prior experimental driving studies with similar objectives. Specifically, an experimental study on hypoglycaemia detection (ClinicalTrials.gov ID: NCT04035993) identified a minimum number of 20 participants [48], while studies analysing alcohol-related effects on driving behaviour typically included sample sizes ranging from 20 to 100 participants [49, 50]. Building on this, an established methodology for estimating model discrimination performance as a function of sample size [51] was applied to data generated in a preceding alcohol-intoxication study [33]. Using this approach, an AUROC of 0.80 for detecting driving above the World Health Organization (WHO)-recommended blood alcohol concentration (BAC) threshold of 0.05 g/dL in intra-individual comparison was projected for a sample size of *n* = 25. Based on previous driving studies by our research group (ClinicalTrials.gov ID: NCT05796609, NCT04980846), a dropout rate of 25% was assumed to account for participant withdrawal (10%), external factors such as weather conditions or test track availability (10%), and expected technical issues (5%) [33, 34, 52]. This results in a target intervention group size of *n* = 33. Driving data from a reference group will be used to evaluate model discriminative power and optimize detection thresholds. Prior hypoglycaemia- and alcohol-impaired driving studies conducted by the same research group indicate that *n* = 10 participants are sufficient for this purpose [33, 52]. To accommodate potential dropout and technical failure, we increased the reference/placebo group size to *n* = 12. Hence, in total, the study targets 45 participants, 33 in the intervention and 12 in the control group.

#### Recruitment and selection

Participants will be recruited through targeted outreach and public dissemination. The recruitment will be supported through printed materials at the University of Bern, online advertisements on relevant social media platforms, and email announcements to students of the University of Bern, the University of St. Gallen, and ETH Zürich, as well as through newsletters of the Automobile Club of Switzerland (ACS), Touring Club Switzerland (TCS), and the Swiss Association for Transport and Environment (VCS). In addition, the recruitment will involve informing participants of the SCRIPT study (“The Safer Cannabis – Research In Pharmacies randomized controlled Trial”, ClinicalTrials.gov ID: NCT06120855) via direct contact and through participating pharmacies distributing flyers. Interested individuals may contact the study team by email or telephone for further information and screening. Reminder emails or telephone calls may be used to support attendance and minimise loss to follow-up. Individuals in a dependent relationship with the study team will not be recruited. These strategies aim to facilitate timely enrolment of the planned sample of 45 participants.

#### Randomisation and blinding

After eligibility is confirmed at the on-site screening (Visit 1), participants are randomly allocated to the intervention group (*n* = 33) or the reference group (*n* = 12) using concealed, pre-randomised allocation cards drawn one at a time. Allocation is performed by a member of the study team who is not involved in the on-site screening procedures, and the allocation cards are not accessible to screening personnel prior to assignment. Recruitment will be monitored to support balanced male/female representation in both study groups, while maintaining the predefined lot-based random allocation. The group assignment is disclosed at the beginning of the study day (Visit 2). The study is conducted open-label and without placebo control, as acute THC administration produces reliably perceptible psychoactive effects that compromise blinding integrity [20, 53]. Therefore, participants, study personnel, and outcome assessors are not blinded to group assignment. Participants are aware of potential cannabis-related impairment during driving, but remain blinded to their measured THC concentrations and the results of performance test assessments conducted before each driving session.

### Study procedures

The study procedures, depicted in detail in SPIRIT Fig. 1, comprise three sequential stages: (i) telephone screening, (ii) on-site screening (Visit 1), and (iii) the controlled driving assessment on the test track (study day, Visit 2).

#### Telephone screening

Telephone screening is conducted up to one month prior to Visit 1. During this call, interested individuals receive detailed information about the study procedures and eligibility requirements. Major inclusion and exclusion criteria are assessed using a structured screening questionnaire, with a focus on medical history and substance use patterns. Cannabis and alcohol consumption habits are evaluated using the CUDIT-R and AUDIT questionnaires, complemented by cannabis-specific screening questions.

Eligible candidates are assigned a unique participant identification number and invited to attend the on-site screening visit.

#### On-site screening (Visit 1)

Visit 1 takes place between two weeks and one month before the driving assessment (Visit 2). Upon arrival, participants are given the opportunity to ask questions before providing written informed consent. Participants then complete questionnaires on driving experience and recent driving behaviour. Wearable sensors (such as Garmin vivoactive 4S) are fitted to enable recording of vital signals during subsequent assessments. Body weight (kg) is measured to determine the individual THC dose for administration at Visit 2. Traffic-medical, traffic-psychological, newly-developed pre-driving performance assessment, and pupillographic tests are administered to establish baseline functional performance. Participants next provide biological samples, including capillary blood for analysis of phosphatidylethanol (PEth), hematocrit, and cannabinoid concentrations; oral fluid and breath samples for cannabinoid analysis; a breath alcohol test; and urine samples for drug screening and pregnancy testing in female participants. These assessments serve to confirm eligibility (e.g., absence of alcohol or other drug misuse) and to establish baseline cannabinoid measures. Following completion of Visit 1, participants are randomly allocated to either the intervention or reference group.

#### Driving assessment (study day; Visit 2)

Visit 2 is conducted as a full-day assessment on a secured, closed test track. Participants are instructed not to consume cannabis or alcohol on the day before, to sleep at least 8 hours during the preceding night, and to consume breakfast as usual. Participants arrive in the morning; a breath alcohol test is performed to confirm sobriety prior to study start, and female participants undergo a urine pregnancy test. Participants also complete questionnaires on recent sleep behaviour and food intake.

A familiarization driving session is conducted before the main study procedure to ensure safe handling of the study vehicle (2020 Volkswagen Touran 1.5 TSI, 110 kW, automatic transmission, 7-seat configuration) and familiarity with the driving procedures and environment. All participants then complete one baseline sober driving session, followed by three additional standardized driving sessions conducted at predefined time windows (approximately 1–2, 3–4, and 5–6 hours after cannabis administration in the intervention group), capturing different levels of cannabis-related impairment over time.

Each driving session consists of three scenarios lasting approximately 10–12 minutes each, representing countryside, urban, and highway driving. These scenarios are designed to expose participants to typical driving demands, such as dynamically changing conditions in urban environments and sustained higher-speed driving on highways. Short breaks of 2–3 minutes are included between scenarios, resulting in a total session duration of approximately 40–50 minutes. All driving is performed on the same test track using a dual-pedal vehicle with a certified driving instructor seated in the front passenger seat, allowing immediate intervention if required. The reference group completes an identical driving schedule, consisting of a sober and three additional driving sessions, without cannabis exposure.

Between driving sessions, structured breaks in both groups are used for recovery, repeated biological sampling (capillary blood, oral fluid, and breath samples), and traffic-medical, traffic-psychological, physiological, and pre-driving test assessments. Participants also complete repeated self-ratings of sleepiness and, in the intervention group, subjective intoxication levels (“feeling high”). These repeated measurements allow characterization of time-dependent changes in driving behaviour, driver state, subjective effects, and biomarker levels. At the end of the study day, assistance with transportation is provided upon request.

#### Intervention: controlled inhalative cannabis administration

Our study adopts controlled inhalative cannabis administration by smoking a prepared joint, as previously applied in experimental THC research [15]. Participants in the intervention group receive a single dose targeting 0.67 mg THC per kg body weight, achieved using THC-dominant cannabis flowers with a confirmed THC content of 15–18%, obtained from Pure Production AG (Laufenburg, Switzerland). A predefined amount of cannabis flowers is mixed with an equal amount of tobacco and rolled into a joint with an additional cigarette filter. The joint is prepared and administrated by trained study personnel, and marked to indicate the point at which the individual target dose is reached. Participants consume the joint under direct supervision by study personnel, who monitor adherence to the standardized smoking procedure (2-second inhalation, 5-second breath-hold, followed by exhalation, with approximately 30 seconds between inhalations) until the marked dose point is reached.

Completion of joint consumption defines time zero for all subsequent assessments and driving sessions. No further cannabis is administered during the study day.

Potential risks include transient cannabis-related effects such as concentration problems, deficits in cognitive perception and in coordination, slowliness/prolonged reaction time, and nausea. These risks are mitigated through strict eligibility criteria, medical and psychological screening, supervised administration, continuous safety monitoring, and driving exclusively on a secured test track in a dual-control vehicle with a certified driving instructor present at all times to enable immediate intervention if required.

There is no direct clinical benefit to participants. Participants may withdraw the study at any time, either by their own decision or by the decision of the study team (e.g., in case of medical concerns, adverse events, or other contraindications to continued participation), with no consequences.

## Data collection and management

### Data collection

Data are collected across all study stages, including telephone screening, on-site screening (Visit 1), and the driving assessment (Visit 2). The study involves multimodal data acquisition from clinical, behavioural, physiological, biological, and in-vehicle sources. Collected data include participant screening and eligibility information; demographic and driving history data; standardized questionnaires assessing substance use (AUDIT, CUDIT-R), sleep behaviour, food intake; results of traffic-medical, traffic-psychological, pupillographic, and pre-driving performance tests; and records of adverse events. The driving is captured using in-vehicle sensor data, including CAN signals and other on-board vehicle sensors. Driver state and behaviour are recorded using DMC, wearable sensors, and additional physiological sensors. Biological samples, including capillary blood, oral fluid, breath samples, and urine, are collected at predefined time points for laboratory analyses of cannabinoids and selected biomarkers. Perceived sleepiness and intoxication levels are also recorded. All data are collected according to standardized procedures defined in the ethics-approved study protocol and are time-referenced to enable synchronization and integrated analysis across data modalities.

### Pseudonymisation and confidentiality

Upon enrolment, each participant is assigned a unique study identification number. This identifier is used consistently across all data sources to ensure effective data integration. A subject identification list (detailing e.g., name or contact details) is stored separately from study data in a secure, access-restricted file. Study data are pseudonymised at the point of collection. The linkage key between participant identification list and study identifiers is stored separately and is accessible only to authorized study personnel. Data used for analysis and machine-learning model development do not contain directly identifiable personal information.

### Data storage and access control

All study data are stored and managed exclusively using the BioMed-IT Node of Leonhard Med (LeoMed), a secure, high-performance research data infrastructure operated by the ETH Zürich and compliant with Swiss data protection regulations (ClinO, Art. 18). Electronic study data, including questionnaires, clinical and psychological assessments, sensor recordings, and laboratory results, are stored in access-restricted directories and are kept strictly separate from the participant identification list. Handwritten source documents, including paper-based case report forms and documentation related to the controlled cannabis administration, are digitized and uploaded to LeoMed. Original paper records are retained in secure institutional storage. Data generated by the study vehicle are initially recorded on local storage for technical reasons, coded on site, and later transferred to LeoMed. Access to raw and processed study data is restricted to authorized members of the study team and limited to purposes consistent with the approved study protocol. All data handling follows applicable institutional guidelines and best practices for information security. Biological samples are identified exclusively by a unique participant number. Samples are collected during the on-site screening (Visit 1) or the driving assessment (Visit 2) and are stored and analysed in restricted-access facilities at IRM, University of Bern. Where specific analyses are subcontracted, samples are transferred securely to external laboratories under contractual arrangements ensuring restricted access and appropriate handling.

### Data quality assurance

Standard operating procedures are used to ensure data completeness, consistency, and integrity. Data are reviewed for plausibility and completeness, and technical checks are performed to identify missing, corrupted, or implausible values. Deviations from planned data collection procedures and technical issues are documented and considered during data processing and analysis.

### Data retention

All study data are archived for at least 20 years after study completion or premature termination. Electronic data, including laboratory results and electronic case report forms, are stored on ETH Zürich servers, while paper-based documents are securely retained at the IRM, University of Bern. Biological samples are analysed at the IRM or designated accredited external laboratories and are destroyed six months after completion of analyses.

### Data and code availability

In accordance with the ethics-approved study protocol (BASEC ID: 2025-01590; Supporting File S2) and applicable Swiss legal data protection requirements, the complete dataset will not be shared. A carefully selected, fully anonymised subset may be made available upon reasonable request and subject to legal, ethical, and consent-based restrictions. Data sharing serves scientific transparency and reproducibility and may support further research and technical development of systems for detecting driving impairment in academic and industrial contexts. The study team derives no financial benefit from such data sharing. All shared data will be anonymised by removal of identifiers and replacement with randomly generated IDs. Raw video and audio data, as well as any other information that cannot be fully anonymised, will not be shared. Prior to any transfer, the specific subset to be shared, the anonymisation procedures applied, and the associated residual re-identification risk will be documented and, where required, submitted to the responsible ethics committee.

Requests for access to anonymised individual participant data should be addressed to the corresponding author. Statistical analysis code used for the evaluation of primary and secondary outcomes will be made available as supplementary material accompanying resulting scientific publications.

### Outcome measures

Based on the assumption that driving behaviour differs between non-impaired and cannabis-impaired states, including several hours after consumption, REVELIO study will leverage differences in driving patterns between intoxicated and non-intoxicated conditions to develop machine-learning models for the detection of cannabis-impaired driving. While machine-learning model performance will establish the overall feasibility of cannabis-impairment detection (primary outcome), the investigation of modality-specific performance and the quantification of changes in relevant behavioural, physiological, and biological metrics (secondary outcomes) will generate new insights and inform the functional and technical requirements for an in-vehicle cannabis-impairment detection system.

#### Primary outcome

The primary outcome is the diagnostic performance of machine-learning models for detecting cannabis-impaired driving using multimodal in-vehicle sensor data, including vehicle CAN data, DMC data, and physiological signals. Model performance will be quantified primarily using AUROC, with accuracy, sensitivity, specificity, and precision as supporting metrics. Models will be trained using labelled data from both sober (baseline) and post-cannabis driving sessions, with impairment detection performance evaluated using session-specific analyses at approximately 1–2, 3–4, and 5–6 hours after cannabis administration. Model performance will be evaluated at both the aggregate, population level and the subject-specific level to assess generalisability and inter-individual variability.

#### Secondary outcomes

Secondary outcomes comprise two domains: (1) diagnostic performance of modality-specific machine-learning models using individual data streams (i.e., CAN-only, DMC-only, and physiological-only data), and (2) deviations from sober baseline in measured behavioural, physiological, performance-related, and biological variables.

Deviation-based outcomes include changes in CAN-derived driving behaviour (e.g., steering, braking, acceleration, and speed-related measures); changes in DMC-derived gaze and head-movement behaviour; changes in driving-related physiological parameters (including heart rate, heart-rate variability, oxygen saturation, and respiration-related signals); changes in instructor-based driving performance assessments and the frequency of safety interventions; changes in performance of in-vehicle readiness and standardized psychometric tests assessing attention and psychomotor function; changes in self-reported subjective intoxication and sleepiness; changes in cannabinoid biomarker concentrations in capillary blood, oral fluid, and breath; and incidence of adverse events.

Performance-related secondary outcomes will be evaluated using the same analytical framework as the primary outcome. Deviation-based secondary outcomes will be evaluated by quantifying changes relative to sober baseline using suitable distributional characteristics (e.g., differences in mean/median/quantiles, variability metrics, event counts, or proportions) as appropriate. All secondary outcomes are assessed at baseline and across three post-consumption driving sessions (approximately 1–2, 3–4, and 5–6 hours after cannabis administration), allowing for both within-subject (baseline vs post-cannabis) and group-wise (cannabis vs reference arm) comparisons.

### Safety considerations

The study is conducted in compliance with the Declaration of Helsinki, International Council for Harmonisation – Good Clinical Practice (ICH-GCP), the Swiss Human Research Act (HRA), and all applicable local regulations, including the Ordinance on Clinical Trials in Human Research (ClinO). Controlled cannabis administration is restricted to a preselected population of recreational cannabis users. Recreational use is defined as self-reported cannabis consumption more than once per month and a CUDIT-R total score *≤*7. Participants with cannabis abstinence (CUDIT-R item 1 = 0) or hazardous use (CUDIT-R *>*7) are excluded, as are vulnerable populations. This threshold ensures prior familiarity with cannabis while reducing the risk of acute adverse reactions in naïve individuals and minimising altered response profiles associated with excessive or dependent use. Participant safety is further ensured through structured screening procedures, including medical eligibility assessment and mandatory pregnancy testing in female participants.

Adverse events (AEs) are defined as any untoward medical occurrences during the study, irrespective of causality. Serious adverse events (SAEs), as defined by ClinO (Art. 63), include life-threatening events, hospitalisation, significant disability, congenital anomaly, or death. Harms are assessed through structured medical monitoring and direct supervision, with additional spontaneous reporting by participants. Causality and severity are evaluated by the investigator and sponsor. SAEs are reported to the sponsor within 24 hours and, if at least possibly related to the intervention, to the responsible ethics committee within 15 days. Ongoing events are followed until resolution or stabilisation.

Biological sampling procedures (capillary blood, oral fluid, breath, and urine) are minimally invasive and associated with low risk. All driving sessions are conducted on a secured, closed test track using a dual-control vehicle, with a certified driving instructor present at all times to enable immediate intervention if required. Comprehensive emergency procedures and insurance coverage are in place.

Participants may withdraw from the study at any time without consequences. While the effects of cannabis administration cannot be reversed once the joint has been consumed, all subsequent study procedures may be discontinued immediately at the participant’s request or for safety reasons. Participants are continuously monitored by trained study personnel, and appropriate medical evaluation or intervention will be initiated if clinically indicated. In the event of early discontinuation, data collected up to the time of withdrawal will be retained and included in analyses unless the participant requests deletion in accordance with applicable data protection regulations.

The study is covered by liability insurance (Baloise) for risk category B in accordance with the ClinO, covering potential trial-related harm for up to 20 years after study completion.

### Statistical analysis

The primary outcome, the diagnostic performance of machine-learning–based detection of cannabis-impaired driving, will rely on supervised classification using multimodal in-vehicle sensor data, including CAN and DMC data. Logistic regression with LASSO regularisation will be used as an interpretable baseline model, reflecting prior evidence of robust performance in detection of driving impairment [33, 48]. This baseline model will be compared with more flexible machine-learning approaches capable of modelling non-linearities and higher-order interactions, such as interpretable Bayesian networks and more data-driven classifiers including Random Forests, Support Vector Machines, or Gradient Boosting models, all operating on engineered predictors derived from sensor data (e.g., steering wheel variability, distance to road edges, gaze dynamics, and eye-movement frequency). Given the high dimensionality of the data, both regularisation techniques (e.g., LASSO) and dimensionality-reduction pre-processing methods (e.g., principal component analysis) will be applied. In parallel, deep-learning approaches, such as convolutional and recurrent neural networks, will be employed due to their potential to learn feature representations directly from the raw data and to capture complex temporal and multimodal patterns in driving behaviour. More broadly, the analysis framework allows for the application of modern artificial intelligence methods that can leverage large-scale multimodal sensor data and improve model generalisation across individuals and driving contexts as methodological advances emerge. Model validation and generalisability will be assessed using strictly participant-level data splits to prevent information leakage. Where computationally feasible given the large-scale, high-frequency sensor data, hyperparameter tuning will be conducted within a nested cross-validation framework. Additionally, subject-level permutation testing will be applied, where feasible, to assess the statistical significance of model performance and determine whether discrimination exceeds chance. This framework will be applied to all secondary outcomes related to cannabis impairment detection across individual data modalities.

For evaluation of secondary outcomes related to changes from sober baseline in behavioural, physiological, performance-related, and biological variables measured during driving sessions, classical statistical inference methods will be applied. These include parametric and non-parametric tests (e.g. t-tests and Wilcoxon tests) for pairwise comparisons, analysis of variance (ANOVA), and (generalised) mixed-effects models to account for repeated measurements within subjects. Mixed-effects modelling will enable adjustment for within-subject correlations and inclusion of time as a structured factor, allowing modelling of temporal effects and capturing the dynamics of outcome changes across post-consumption driving sessions relative to baseline. Analyses will include both within-subject comparisons (post-consumption vs baseline) and group-wise comparisons (cannabis vs reference group), including evaluations conditional on subjects’ characteristics (e.g., demographics, cannabis-use frequency).

Analyses will primarily follow a per-protocol approach. Sensitivity analyses may include all randomised participants with available data.

### Ethical considerations

The study is conducted in accordance with the Declaration of Helsinki, ICH-GCP, HRA, and ClinO. Ethical approval was granted by the Cantonal Ethics Committee Bern (BASEC ID: 2025-01590), subject to the authorization from the Swiss Federal Office of Public Health (BAG) for the use and administration of cannabis in the study intervention group. The trial was classified as risk category B due to the controlled administration of cannabis to pre-screened recreational users.

Written informed consent is obtained in person by the authorised study team member prior to any study-specific procedures. Participants may withdraw from the study at any time without disadvantage. Only healthy adult recreational cannabis users are enrolled in the study; no vulnerable populations are included. Risks are minimised through eligibility criteria, medical and psychological screening, pregnancy and breath alcohol testing, continuous safety monitoring, and driving exclusively on a closed test track using a dual-pedal vehicle with a certified driving instructor present at all times. Biological sampling procedures are minimally invasive. All analyses and system development are conducted post hoc. No automated driving restrictions, system-initiated interventions, or legal consequences are implemented, and the study does not replace established medical or legal assessments.

Participants receive fixed compensation of CHF 30 for the on-site screening visit (Visit 1) and CHF 220 for the driving assessment (Visit 2), independent of study completion. Confidentiality and data protection are ensured through pseudonymisation, restricted access, and secure data storage in accordance with Swiss data protection regulations.

#### Sponsor, trial governance and monitoring

The University of Bern serves as the trial sponsor and coordinating site. The responsible party and principal investigator (PI) is Prof. Dr. Wolfgang Weinmann, IRM, University of Bern, Switzerland. Overall scientific and operational responsibility lies with the PI and the academic study team.

Given the single-centre design and pilot nature of the trial, no independent steering committee, endpoint adjudication committee, or data monitoring committee has been established. No interim analyses or formal stopping rules are planned. Data management and quality control are performed in accordance with institutional and regulatory requirements. Trial conduct and data quality are monitored by an independent internal monitor (PD Dr. `es sc., MLaw Martin Zieger, IRM, University of Bern), who is not involved in participant enrolment or intervention procedures.

Monitoring includes verification of informed consent, eligibility criteria, adherence to protocol procedures, correct group allocation, and data completeness. An initial review is conducted after the first two participants, with additional reviews performed as required. Identified issues are reported to the PI within 24 hours and corrective actions are implemented.

#### Patient and public involvement

Patients or members of the public were not involved in the design or conduct of this study. Recruitment may include outreach to individuals participating in existing cannabis-related research studies, who meet the eligibility criteria. No formal patient or public involvement informed the study.

#### Study status and timeline

The study received initial ethical approval from the Cantonal Ethics Committee Bern (BASEC ID: 2025-01590) on 28 November 2025. The final BAG authorisation for cannabis distribution was received on 24 March 2026. The amended study protocol (version 5.1) has been approved and is provided as Supporting Information S2. The statistical analysis plan is incorporated within the approved protocol (pages 34–35) and the present manuscript. The study is registered at ClinicalTrials.gov (NCT07401628). Registration was submitted on 22 January 2026 following ethics approval and prior to enrolment of the first participant; the record was posted on 10 February 2026 after administrative review. Recruitment activities (advertisement) have begun on 5 January 2026 and remain ongoing at the time of manuscript submission. The first on-site screening visit (Visit 1) was conducted on 9 February 2026, and the enrolment procedures are ongoing. The first driving assessment (Visit 2) on the closed test track was conducted on 24 March 2026, and the completion of data collection is anticipated by 19 June 2026. Any protocol amendments will be submitted to the responsible ethics committee and updated in the trial registry prior to study implementation. The approved amendments will be communicated to investigators and, where relevant, to trial participants. In the event of premature termination, the ethics committee and trial registry will be informed in accordance with applicable regulatory requirements.

## Discussion

This manuscript presents the REVELIO study protocol, a randomised, controlled, single-centre pilot trial designed to generate synchronised multimodal in-vehicle data under controlled cannabis exposure. The primary objective is to develop and test the feasibility of machine-learning–based distinguishing sober from cannabis-influenced driving states using vehicle CAN data, DMC outputs, and physiological signals. By combining standardized exposure conditions with real-vehicle operation, the study establishes a structured experimental framework for assessing the feasibility and diagnostic performance of data-driven cannabis impairment detection.

The planned sample size of 45 participants (33 intervention, 12 reference) reflects an exploratory pilot design with the intention of balanced sex representation. While sufficient for model training and internal cross-validation under controlled conditions, the sample size limits heterogeneity across age, cannabis-use patterns, and behavioural variability of study participants. Furthermore, the use of THC-dominant cannabis with a defined concentration range standardizes exposure but does not capture the full diversity of cannabinoid profiles encountered in real-world settings, such as including cultivars with higher cannabidiol (CBD) content or varying terpene compositions, whose effect may vary [54]. Larger and more heterogeneous cohorts will therefore be required in the future to better evaluate robustness and generalisability of study findings.

A central feature of the protocol is the integration of multimodal in-vehicle data with synchronous biological and functional reference measures. Cannabinoid concentrations in capillary blood, oral fluid, and breath serve as objective exposure markers enabling to assess pharmacokinetic profiling of biomarkers, while traffic-medical, cognitive, and psychomotor assessments enable functional characterisation of driver’s performance and state over time. This design allows examination of the relationship between biological concentrations and observable performance- and functional-related patterns of subjects driving a real vehicle, without relying solely on substance detection. Driving is conducted in a dual-control vehicle on a secured test track to ensure safety and standardization. While investigation in a controlled environment represents an essential first step, further research in larger and more heterogeneous populations under real-world traffic conditions will be necessary to extend these findings, establish external validity, and ensure transferability to complex public road environments.

The present study does not implement automated driving restrictions, system-initiated vehicle interventions, or legal consequences. The developed models are evaluated in a research context only and do not replace established medical or legal assessments, nor do they introduce de facto driving eligibility determinations for study participants.

Importantly, the protocol is methodologically aligned with prior controlled driving studies conducted by the same research group investigating detection of alcohol- and hypoglycaemia-related driving impairment (ClinicalTrials.gov IDs: NCT04980846, NCT05796609, NCT04035993), using the same study vehicle, driving procedures, and sensor configuration. This methodological alignment allows for pooled integration of existing multimodal datasets with those from REVELIO, enabling subsequent cross-condition analyses. These analyses will examine whether shared functional impairment patterns can be identified across different physiological and pharmacological states and will explore the feasibility of a general, cause-independent “fit-to-drive” detection framework focused on overall driving impairment rather than specific substances or medical conditions.

In summary, the REVELIO pilot test-track trial provides a multimodal dataset for the development and initial evaluation of a real-time, in-vehicle approach to detect cannabis-related driving impairment. As an exploratory study, it is designed to assess feasibility and model performance under controlled conditions. Although further validation in larger and more diverse populations will be required before considering real-world implementation, the conceptual and methodological alignment with related alcohol- and hypoglycaemia-focused studies establishes a consistent framework for subsequent cross-condition analyses and exploration of a broader, cause-independent fit-to-drive detection approach.

Study findings will be disseminated through peer-reviewed publications, scientific conferences, and reporting in the ClinicalTrials.gov registry.

## Data Availability

In accordance with the ethics-approved study protocol (BASEC ID: 2025-01590) and applicable Swiss legal data protection requirements, the complete dataset will not be shared. A carefully selected, fully anonymised subset may be made available upon reasonable request and subject to legal, ethical, and consent-based restrictions. Data sharing serves scientific transparency and reproducibility and may support further research and technical development of systems for detecting driving impairment in academic and industrial contexts. The study team derives no financial benefit from such data sharing. All shared data will be anonymised by removal of identifiers and replacement with randomly generated IDs. Raw video and audio data, as well as any other information that cannot be fully anonymised, will not be shared. Prior to any transfer, the specific subset to be shared, the anonymisation procedures applied, and the associated residual re-identification risk will be documented and, where required, submitted to the responsible ethics committee.

## Funding

This project is primarily funded by the Fonds für Verkehrssicherheit (FVS), a Swiss public institution (grant number 22.25.01), covering 84% of the total project costs. The remaining 16% are covered by the University of Bern, ETH Zürich, and the University of St. Gallen through institutional contributions, including personnel time of the project leaders, doctoral salaries, and infrastructure-related expenses such as IT support, analytical chemistry, and data processing. The University of Bern acts as sponsor and is responsible for overall study conduct in accordance with Swiss regulatory requirements. Scientific design, conduct, analysis, and reporting of the study are under the responsibility of the academic investigators. The FVS had no role in the study design or in the preparation of this manuscript and will have no role in data collection, data analysis, or interpretation of study results. Publication is subject to formal approval in accordance with the FVS funding regulations.

## Competing Interests

The authors declare no competing interests.

## Acknowledgements

M.J. is affiliated with the Centre for Digital Health Interventions (CDHI), a joint initiative of the Institute for Implementation Science in Health Care, University of Zurich, the Department of Management, Technology, and Economics at ETH Zurich, and the Institute of Technology Management and School of Medicine at the University of St.Gallen. M.J. acknowledges funding support from CSS, a Swiss Health Insurer. However, CSS was not involved in this study.

## Supporting information

### S1. SPIRIT 2025 Checklist

Completed SPIRIT (Standard Protocol Items: Recommendations for Interventional Trials) checklist for the REVELIO study protocol.

### S2. Ethics-approved study protocol

Full study plan (version 5.1), representing the amended protocol approved by the Cantonal Ethics Committee Bern (BASEC ID: 2025-01590).

## Notes

### Competing Interest Statement

The authors have declared no competing interest.

### Clinical Trial

NCT07401628

### Author Declarations

The study was approved by the Cantonal Ethics Committee Bern, Switzerland (BASEC ID: 2025-01590).

